# Evaluating a PEN-Plus training of trainers program in cardiac point-of-care ultrasound in Sub-Saharan Africa: Protocol and initial results

**DOI:** 10.1101/2025.09.29.25336919

**Authors:** Sheila L Klassen, Alma J Adler, Marta P Rodriguez, Gene F Kwan, Gedeon Ngoga, Catherine Karakezi, Emily B Wroe, Gene Bukhman

**Affiliations:** Center for Integration Science, Division of Global Health Equity, Department of Medicine, Brigham and Women’s Hospital, Boston, USA; Division of Cardiovascular Medicine, Department of Medicine, Brigham and Women’s Hospital, Boston, USA; Program in Global Noncommunicable Disease and Social Change, Department of Global Health and Social Medicine, Harvard Medical School, Boston, Massachusetts, USA; Section of Cardiovascular Medicine, Department of Medicine, Boston University Chobanian and Avedisian School of Medicine, Boston Medical Center, Boston, USA; NCD Alliance Kenya

**Author notes:** Corresponding author: Dr. Sheila Klassen 75 Francis St. Boston MA 02115 USA.

## Abstract

**Introduction:** Access to cardiac diagnostics is limited in rural areas of low- and lower middle-income countries (LMICs). The Package of Essential Noncommunicable Disease Interventions - Plus (PEN-Plus) strategy decentralizes and integrates care for severe noncommunicable diseases, including cardiac disease, to more accessible facilities for this population. Given the shortage of healthcare providers, training frontline clinicians to perform simplified cardiac point-of-care ultrasound (POCUS) and equipping them to train others is essential. This study describes a novel approach to preparing non-specialists as trainers and mentors in cardiac POCUS.

**Methods:** This study evaluates the effectiveness and implementation outcomes of a training of trainers (ToT) program within the PEN-Plus strategy. Midlevel providers and generalists from five African countries were nominated by their employers as candidate trainers. The seven selected candidates participated in virtual and in-person training, which covered essential teaching skills and provided supervised cardiac POCUS teaching simulation. The Kirkpatrick model provided the program evaluation framework. Results of the initial post-training assessment encompassing Level 1 (Reaction) and Level 2 (Learning Assessment) are presented here. Level 3 (Behavior) will be assessed at 6 months and examine skills application, training environment, and barriers. Level 4 (Results) will be evaluated using the RE-AIM implementation framework at 12 months. Exploratory analyses will examine links between initial training assessment variables and short-term implementation outcomes.

**Preliminary results:** Most trainers reported high satisfaction with the ToT program. All strongly agreed that content was relevant and they would recommend the program to colleagues. Experts scored most trainers as Excellent or Very Good in all Level 2 domains. Most trainers attained the highest score across all competencies on trainee evaluations.

## Introduction

In low- and lower middle-income countries (LMICs), cardiac care is delivered predominantly at tertiary care hospitals in large urban areas (1). However, the majority of the population in most LMICs reside in rural areas and receive health care at non-tertiary care facilities (2). These populations also make up the majority of the bottom billion, or poorest billion people in the world (3). Cardiac disease in LMICs often manifests at late stages of heart failure and at a younger age than in high-income countries, with a very poor prognosis (1,4–6). Most of the data generated from LMICs on cardiac disease originate from tertiary care centers (1), making true incidence and prevalence of cardiac disease in LMICs unknown. In rural Rwanda, the majority of cardiac disease is caused by rheumatic heart disease, congenital heart disease, cardiomyopathy, and hypertensive heart disease. In this Rwandan study, the median age at diagnosis is 27 years old (7).

The Package of Essential Noncommunicable Disease Interventions - Plus (PEN-Plus) model of care (8) aims to decentralize and integrate care for severe noncommunicable diseases (NCDs) including cardiac disease to primary referral hospitals. PEN-Plus provides easier access to high-quality care at rural hospitals for impoverished populations (9) and complements the World Health Organization (WHO) Package of Essential Noncommunicable Interventions for Primary Care (PEN), a primary care strategy for non-severe NCDs. Outpatient PEN-Plus clinics at rural district hospitals are staffed by mid-level providers – clinical officers, physician assistants, general practitioners, and trained nurses – who care for patients with severe NCDs including cardiac disease. An essential skill for PEN-Plus providers is cardiac point-of-care ultrasound (POCUS), used to diagnose and manage common cardiac conditions such as cardiomyopathy, rheumatic heart disease, and congenital heart disease (10). Diagnosis and management are guided by PEN-Plus cardiac adult and pediatric protocols (Appendix 1 & 2), which are designed to diagnose and treat cardiac disease most endemic to the bottom billion population (11).

The PEN-Plus strategy was adopted as regional strategy by the WHO African Region in 2022 (12) and is currently implemented in 14 countries (13) by local implementing organizations (Figure 1). WHO African Region is targeting implementation of diagnostic and therapeutic services for severe NCDs in 70% of Member States by 2030 (12). PEN-Plus implementing organizations work in partnership with the Ministries of Health and form part of the NCDI Poverty Network with co-secretariats in Boston, United States and Maputo, Mozambique. During initial implementation, training of PEN-Plus providers in cardiac POCUS is undertaken by local experts such as cardiologists located in the capital city where available. When local cardiologists are not available, experts from the Boston co-secretariat travel internationally to carry out training. The implementation of PEN-Plus is a phased theory of change where, after initial implementation at one or several sites, National Operational Plans are developed in subsequent years for expansion of clinics into other parts of the country. For this expansion, more local trainers are needed.

**Figure 1:**
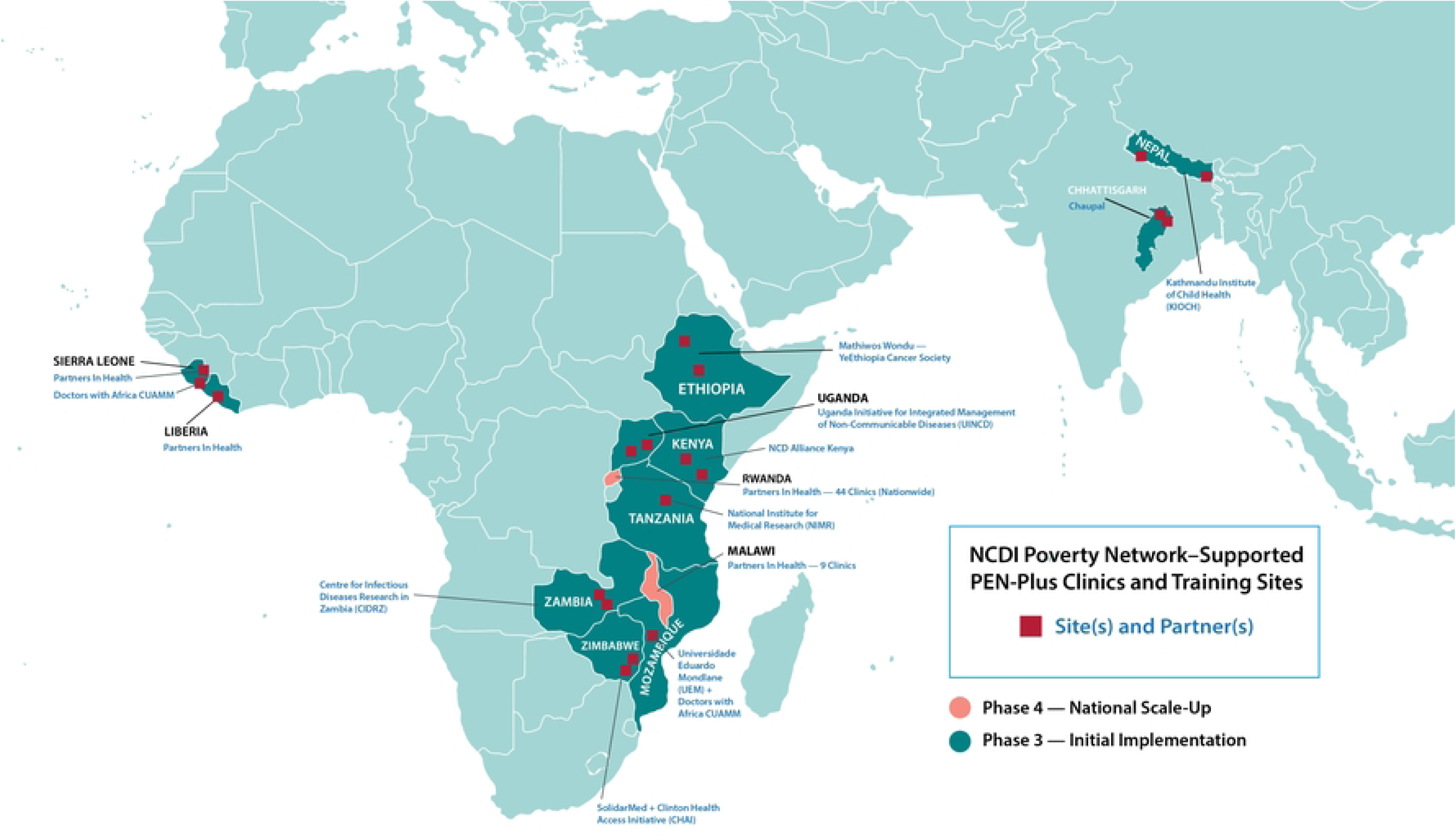
Map of PEN-Plus implementing partners.

Because of the very small number of healthcare providers in LMICs relative to the population and concentration of specialists and experts in urban areas, the peer training model or training of providers who can train others is essential to decentralization.

Programs that aim to train trainers in LMICs exist, especially in places where task-shifting for more specialized knowledge is required (14–16) but only two were skills-based focusing on resuscitation. None covered cardiac disease care or performance of ultrasound. In this study we describe the protocol for a novel method of training healthcare workers to train and mentor other mid-level providers in PEN-Pus cardiac POCUS, and provide the initial results. The aim of this study is to determine the effectiveness and implementation outcomes of a TOT program.

## Methods

### Study design

This is a prospective cohort study of seven PEN-Plus clinicians from five countries in Sub-Saharan Africa who were trained virtually in a ToT program from March 10 to April 11 2025 followed by a one-week in-person training session in Kenya from April 13 to April 17 2025. Clinicians underwent a selection process which incorporated knowledge assessments and programmatic goals. The Kirkpatrick model was used to design training assessments. Preliminary data was gathered immediately following training in Kenya and additional data will be gathered from trainers virtually at 6 months (November 2025) and 12 months (May 2026).

### Frameworks

The Kirkpatrick model (17) is a globally recognized training evaluation model and served as a guide for the creation of evaluation materials for this ToT program (Figure 2). The Kirkpatrick model is commonly used in LMICs (18–21) and provides a structure for gathering both subjective and objective data on training, emphasizing the evaluation of real-world results. It is adaptable to all types of training and incorporates an evaluation of the implementation of training content at some time point after training has taken place. Its four levels provide a framework for detailed and longitudinal training evaluations.

**Figure 2:**
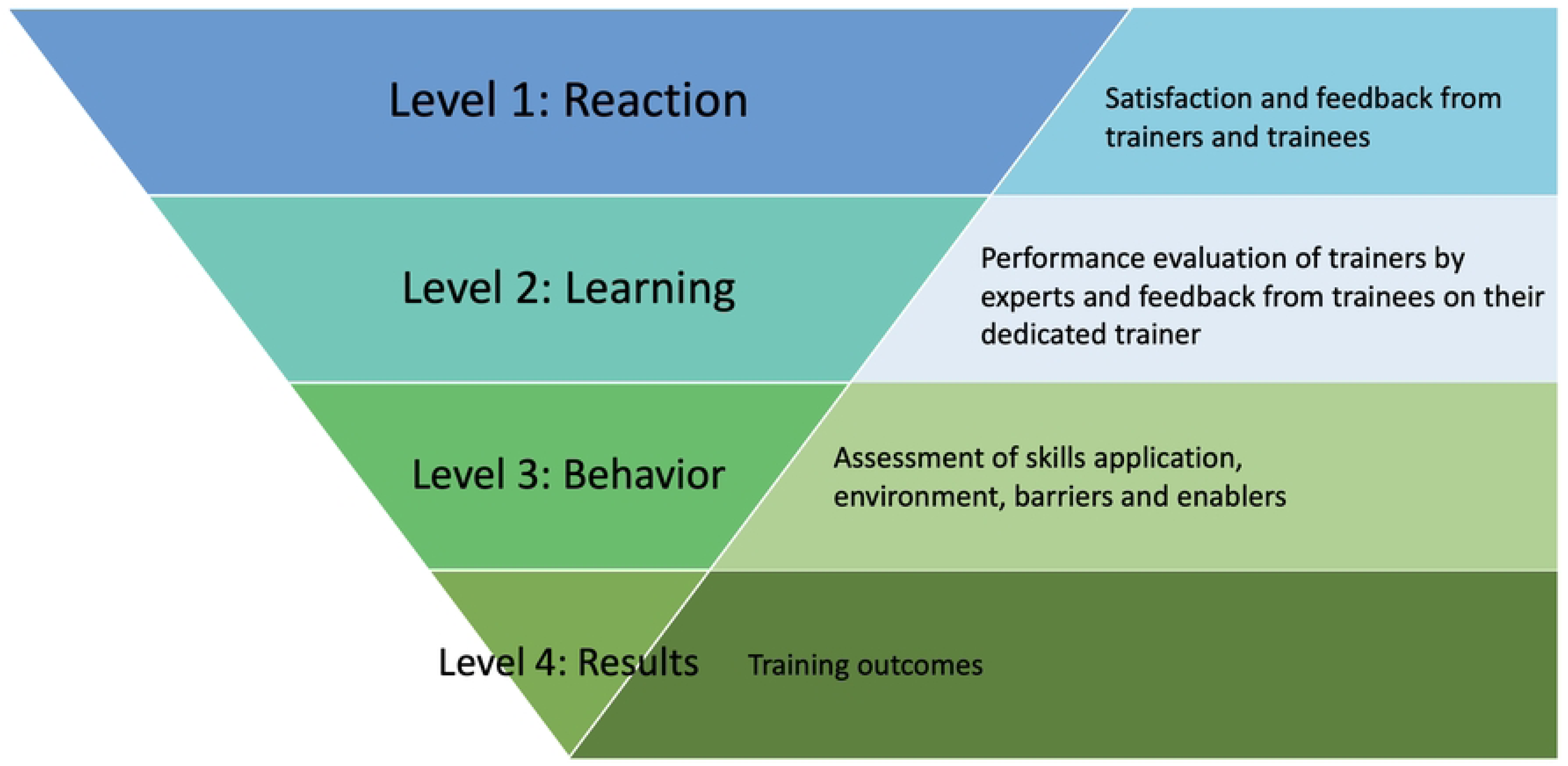
Evaluation of the master trainer training program based on the Kirkpatrick model.

To examine Results in Level 4 of the Kirkpatrick model, we will measure implementation outcomes using the RE-AIM framework (22) at 6- and 12-months post training at clinicians’ home institutions.

### Setting

Five virtual meetings were held once weekly (March 10-April 7, 2025) followed by five days of in-person training (April 13-17, 2025) in a rural referral hospital in Isiolo, Kenya. Follow-up data collection will be performed virtually in November 2025 and May 2026.

## Participants

### Trainer nominations

A request for nominations was put forth to all Phase 3 PEN-Plus implementing organizations in 10 countries (Figure 1) in January 2025 for mid-level providers who were previously trained in PEN-Plus and could potentially assume the role of trainer within their PEN-Plus site.

Requirements for nomination included prior in-person training in cardiac POCUS and an active role in PEN-Plus where they work, a commitment from partner organizations to offer stable employment to the trainer for the next two years and to ensure that training peers locally would be part of their role within the PEN-Plus clinic. This commitment from partners was important to mitigate the problem of staff turnover when investment had been made in a specialized ToT program. Partner organizations were not required to nominate a candidate if they felt the ToT model did not fit with their National Operational Plan or if they did not feel they had a good candidate to offer. Nominees were expected to have a good command of the English language to participate; all training materials were only provided in English.

### Trainer baseline knowledge assessment

Nominees underwent a three-part assessment, which was submitted online, to evaluate their cardiac POCUS skills and ability to use cardiac POCUS for patient diagnosis and management. The purpose of the assessment was to ensure trainers had adequate knowledge and skills so that the ToT program would not involve additional “catch up” medical teaching. First was a 32-question multiple choice quiz (MCQ) on basic cardiac anatomy and ultrasound physics, patient assessment, and management of the common PEN-Plus cardiac conditions found in Appendix 1.

This MCQ exam has been used over the past five years in multiple cohorts of healthcare providers across the NCDI Poverty network (10,23) with participant feedback confirming that the MCQ is easily understood and reflective of the didactic material. The second part of the assessment consisted of an MCQ asking providers to interpret cardiac POCUS images from five patient cases, make a diagnosis based on a case stem and images, and identify the correct management plan. The third part of the assessment required providers to submit five cardiac-focused patient cases from clinical practice at their home institution for expert evaluation. The cardiac POCUS images must have been performed by the nominee and the nominee must have either independently performed or played a key part in the evaluation, diagnosis, and initial management of the patient.

All assessments were created by a cardiologist with seven years of experience in PEN-Plus cardiac training (SLK). The submitted cases were graded by two cardiologists [SLK, GFK] and one internist with medical education expertise (MPR), all with many years of experience training on PEN-Plus. MCQs were given a percentage score and cases were graded on a five-point scale on elements of image quality, correctness of diagnosis, and initial management.

### Trainer selection and study size

Out of the twelve nominees, seven were selected. Of the five nominees who were not accepted, one did not speak English, two withdrew their nominations, one was a physician not employed by the nominating organization and planned to be going for cardiology fellowship training, and one came from an organization who did not intend to use the peer to peer training model for clinic expansion.

Implementing organizations provided written commitment to use the new skills of these nominees in ongoing peer mentorship efforts, training new staff in cases of staff turnover within PEN-Plus clinics, and for the training of new PEN-Plus providers if new clinics are opened.

Because PEN-Plus is a multi-country strategy and previous trainers have traveled to adjacent countries to train providers in new PEN-Plus clinics, the cross-cultural participation of trainers from multiple countries in this ToT program was an intentional decision and a strength of the program.

## Training format

The five virtual 60- to 90-minute meetings (held once weekly) covered pre-training material for the trainers on topics such as how to give feedback, practicing PowerPoint presentation skills, optimization of cardiac POCUS image quality, and to hear lessons learned from an existing experienced PEN-Plus trainer.

Subsequent to virtual training, five days of in-person training took place in a rural Kenyan referral hospital with seven trainers from five different countries, fourteen local rural Kenyan trainees (to match trainer to trainee in a 2:1 ratio), and four experts to supervise the seven groups (Figure 3). Experts consisted of the two cardiologists and one internist who had graded the submitted cases during the assessment phase and as well as a nurse who was an existing experienced PEN-Plus trainer (GN). Program managers and administrative staff were present to coordinate all training participant and patient logistics. The local hosting organization was NCD Alliance Kenya and representatives from the Kenyan Ministry of Health came to observe parts of the ToT program.

**Figure 3:**
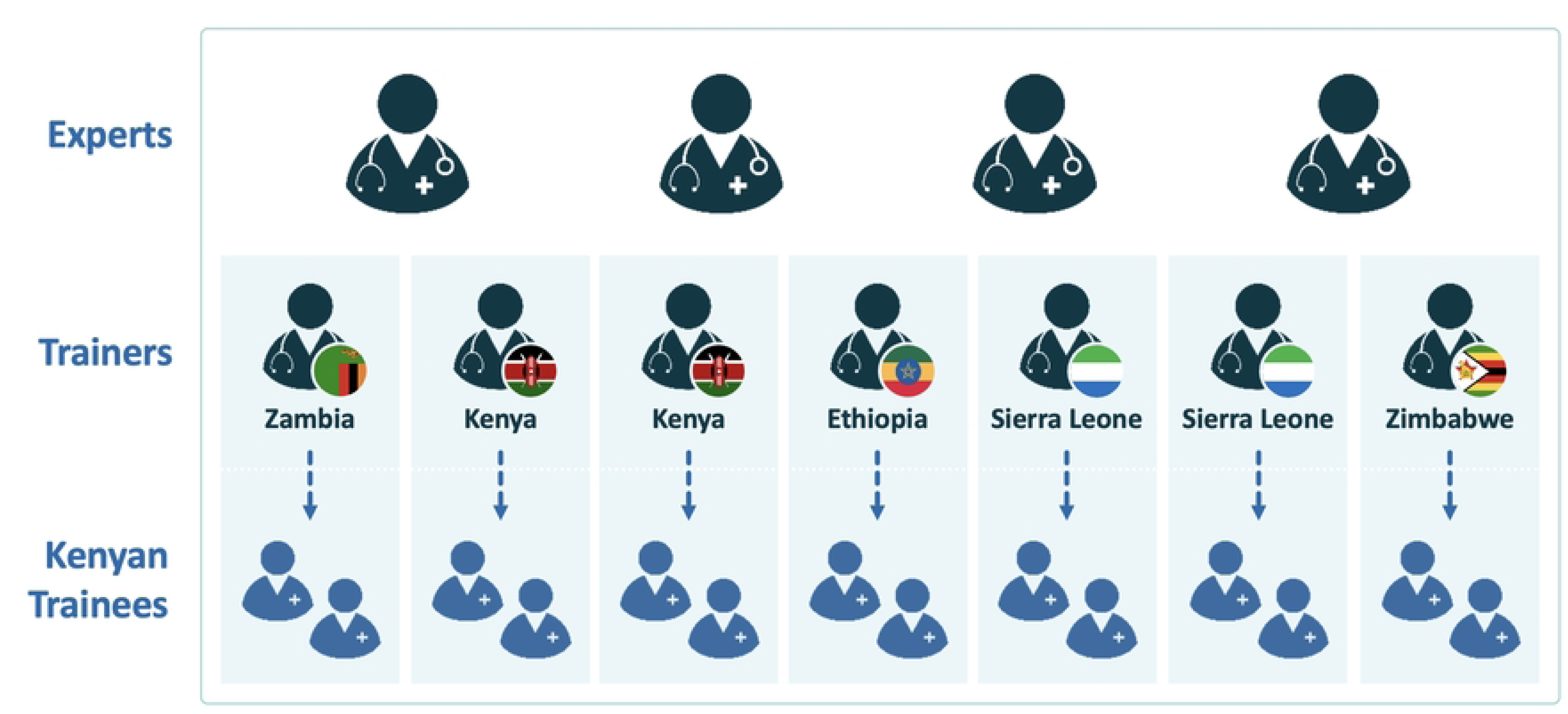
Structure of in-person master trainer program supervision. Trainers were from Ethiopia, Kenya, Zambia, Zimbabwe, and Sierra Leone. International experts were two cardiologists, one internist, and one experienced nurse.

During the in-person training week, trainers delivered a didactic lecture on basic ultrasound physics and cardiac POCUS ultrasound views to trainees. Trainees consisted of nurses and clinical officers who had minimal to no cardiac POCUS or heart failure experience. In small groups, trainers demonstrated how to obtain cardiac POCUS views on a volunteer. All trainees were able to practice all views on a volunteer with assistance from the trainers.

For the rest of the week, the trainer and trainee groups performed scanning practice in the morning, then discussed cases in the afternoon. Afternoon case discussions consisted of image review of interesting patients who had been scanned by various groups in the morning as well as review of patient cases from a PEN-Plus online training case bank. Trainers led case discussions on evaluation of cardiac patients, interpretation of POCUS images, and management of particular diagnoses. Figure 4 describes the virtual and in-person structure and teaching topics of the program. Trainers were encouraged to give constructive feedback to each other and their trainees. All activities were supervised by experts. A daily debrief session was held with trainers and experts for trainers to reflect on the day’s experiences and learn from each other.

**Figure 4:**
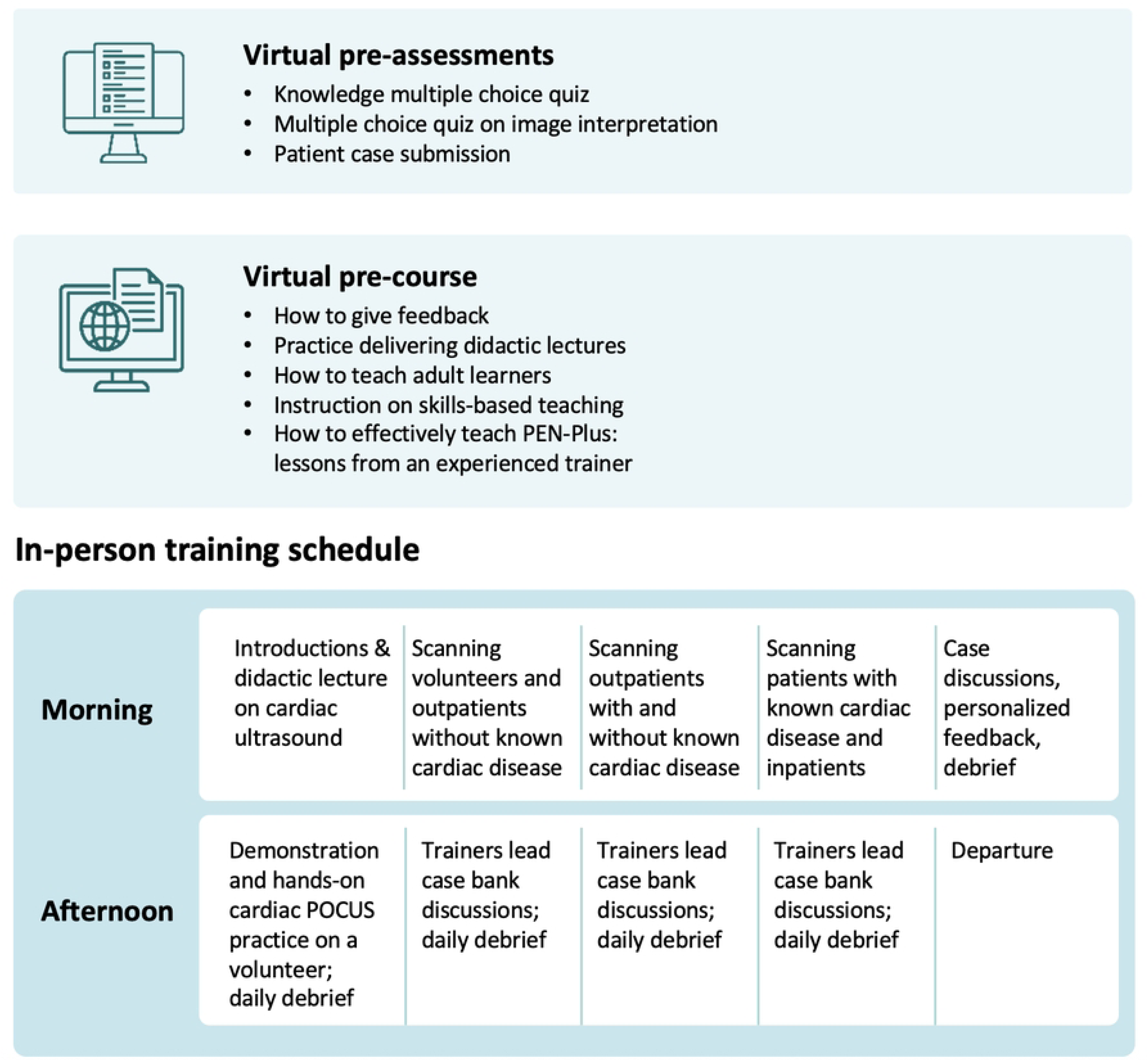
Virtual and in-person components of the program.

Two virtual training calls were held in May 2025 after the in-person training for trainers to debrief and reflect on lessons learned, and to develop training plans within their home PEN-Plus clinics.

The six month post training evaluations (Level 3 and 4) will be administered starting October 15, 2025 and the 12 month evaluation (Level 4 only) will be conducted in May 2026.

Figure 5 depicts the steps of the training program from nomination to evaluation of implementation outcomes.

**Figure 5:**
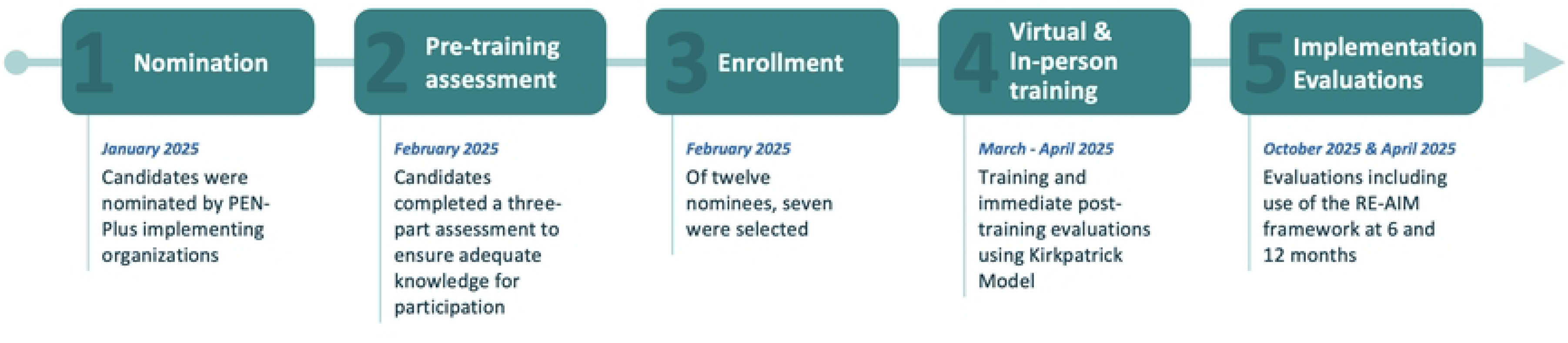
Training program steps from beginning to end

## Evaluation tools and variables

The Kirkpatrick model is used in this study to evaluate the training program.

The initial post-training assessment was completed by trainers, their trainees, and supervising experts with Level 1 (Reaction) evaluating satisfaction with training. Participant satisfaction has been identified as an important component of training, particularly participant perception of usefulness of training (24). In Level 1, trainers were asked to complete an evaluation of the ToT program. Questions in this evaluation asked trainers about the relevance and value of the training program to their work and their impressions on the quality of program design.

Level 2 (Learning) evaluated trainer performance in the domains of Knowledge, Teaching Skills, Communication Skills, Leadership Skills, and Participation and Attitude. In Level 2, Knowledge and Teaching Skills included components considered essential from a PEN-Plus programmatic standpoint. Leadership skills were deemed necessary for trainers to design and conduct cardiac POCUS training at their home facilities and other studies have found that ToT courses have been effective at fostering leaders (25). Level 2 consists primarily of an evaluation of the trainer by experts but incorporates evaluation by trainees on perceived teaching and communication skills of the trainer as well as ability of the trainer to give feedback, an important component of medical education (26). In Level 2, trainers were evaluated by both the experts who were directly supervising their training (Table 2) and the two trainees they taught during the in-person week (Table 3). Because experts were able to observe all trainers during the in-person week, all experts graded all trainers, which incorporates a degree of rigor into trainer assessment.

Level 3 (Behavior) will be completed 6 months after training and is structured according to the categories of Application of Skill, Support and Environment, and Barriers and Enablers. In Level 3, trainers will evaluate the impact of the ToT program on their practice and role as a trainer at their home organization. This will take place 6 months after the in-person week in November 2025.

Full evaluation tools can be found in Appendix 3.

Level 4 (Results) of the Kirkpatrick Model was structured using the RE-AIM framework using the categories of Reach, Effectiveness, Adoption, and Implementation to evaluate actual implementation of training by trainers, and evaluated at the 6 and 12 month time points. This Implementation Assessment is shown in Table 1. The Maintenance category will contain the same questions found in the preceding categories (Reach, Effectiveness, Adoption & Implementation) but responses will be gathered at the 12 month time point. RE-AIM will be completed as one per country rather than individually by the Trainer for programmatic purposes. Trainers will be provided a mentorship checklist to grade competencies of trainees which allows them to track progress over time. The mentorship checklist is a way of helping trainers evaluate their trainees and is not used in this study for data gathering. Given close Network collaborations, including between the study team, trainers, and sending organizations, no loss to follow-up is anticipated.

**Table 1:** Implementation assessment with variables organized according to the RE-AIM framework.

| Category | Variable |
| --- | --- |
| Reach | # of facilities where trainers are actively training relative to<br>a) total number of PEN-Plus facilities in the country<br>b) total number of PEN-Plus facilities in Sub-Saharan Africa<br># facilities where new PEN-Plus clinic openings are made possible by trainers |
| Effectiveness | # of trainees that can appropriately identify patients who need echo performed<br># of trainees (new/existing) that can independently perform echo, scoring 4 or more on image quality on the mentorship checklist.<br># of trainees (new/existing) that can describe echo criteria for cardiac diagnoses included in the PEN-Plus protocol<br># of trainees (new/existing) that can make a correct diagnosis based on echo findings and following the PEN-Plus protocol (score 4 or more on image quality on the mentorship checklist) |
| Adoption | # of PEN-Plus organizations who have implemented training by trainers as a key programmatic component<br># of trainers who are actively carrying out a self-designed training plan |
| Implementation | # of trainees who completed echo training at local facility<br># of days of didactic lectures carried out by the trainer<br># of days per month the trainer spends training new trainees on how to perform echo<br># of days per month the trainer spends mentoring existing trainees<br># Echocardiograms done per trainee with:<br>a) active assistance by a trainer<br>b) no active assistance by a trainer |
| Maintenance | Re-evaluation of Reach, Effectiveness, Adoption, and Implementation categories at 12 months (May 2026) |

## Data sources

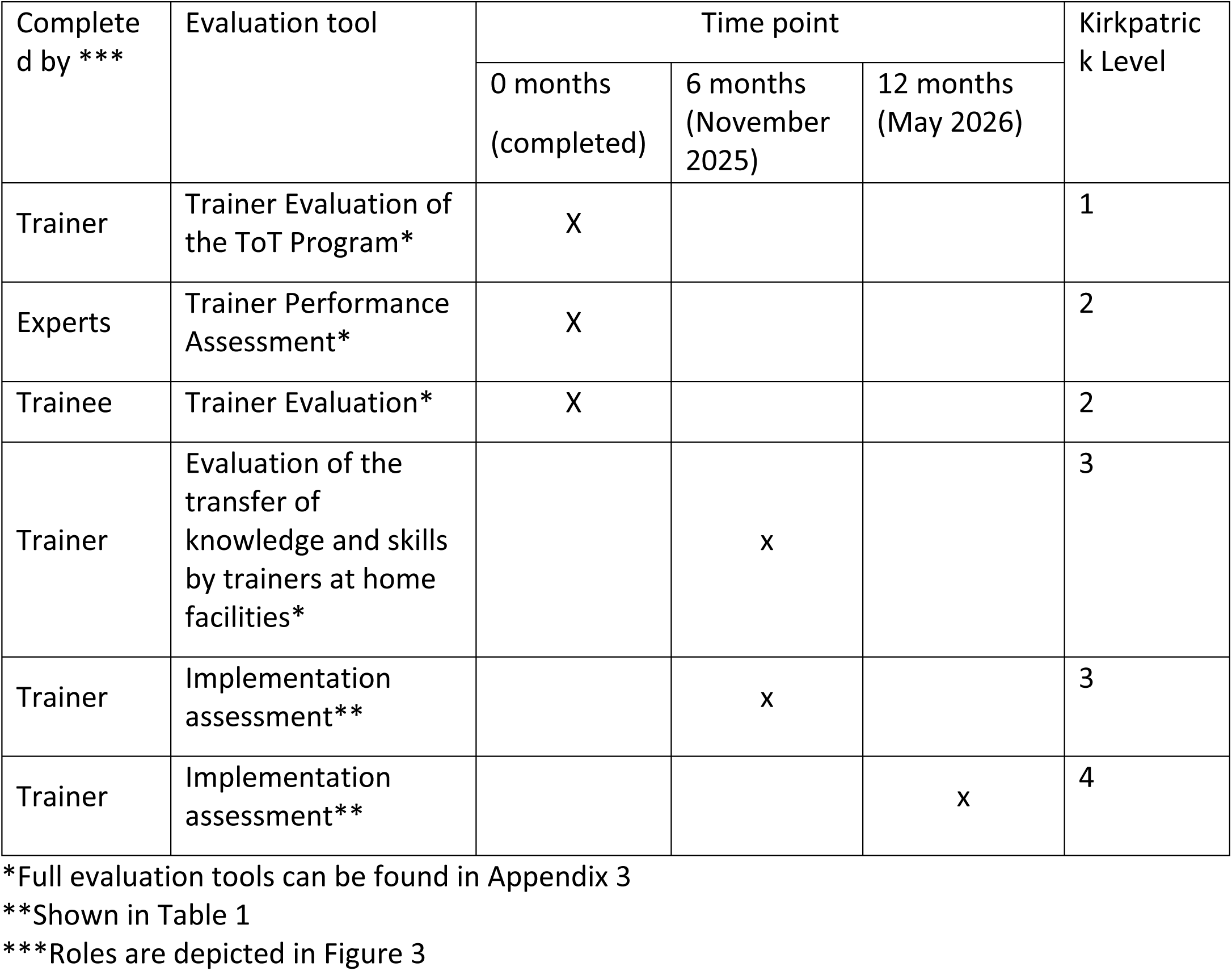

### Ethics and dissemination

An institutional review board application for data analysis is approved through the Brigham and Women’s Hospital (Protocol #: 2025P001302). All trainers were informed that training data would be analyzed and provided implied consent for the study by participating in training. For dissemination, complete results of this study will be submitted for publication in a peer-reviewed journal and via reports to NCDI Poverty Network stakeholders.

## Data Analysis

### Quantitative data

For initial data, quantitative data such as Likert scores were analyzed using descriptive statistics. Categorical variables were examined using frequencies and percentages. Quantitative data analysis was performed using STATA version 17 and Microsoft Excel. The same process will be applied to follow-up data. In follow-up data, continuous variables will be expressed using mean and standard deviation. Correlations between preliminary and follow-up data will use Spearman’s rank correlation.

### Qualitative data

Free text responses in the surveys underwent a thematic content analysis. Responses were grouped into themes. A descriptive definition was provided for each category. The number of unique themes and the frequency of codes within each theme were reported.

## Patient and public involvement

No patients or patient data are involved in the study and data analysis. Collected data was and will be limited to evaluation of the trainers and performance of their trainees.

## Results

### Kirkpatrick Level 1 – trainer satisfaction with training

All trainers strongly agreed that the content was relevant to their work and they would recommend the ToT training to a colleague. Six of the seven trainers strongly agreed that training met their expectations, that training materials were easy to understand, and experts were knowledgeable and effective, while one responded agree for each of those questions. Five trainers strongly agreed that training was engaging and held their interest while two agreed. Four strongly agreed that the training environment was comfortable and conductive to learning while three agreed. For the question “the pace of learning was appropriate”, four trainers strongly agreed, three agreed, and one was neutral. Results are depicted in Figure 6.

**Figure 6:**
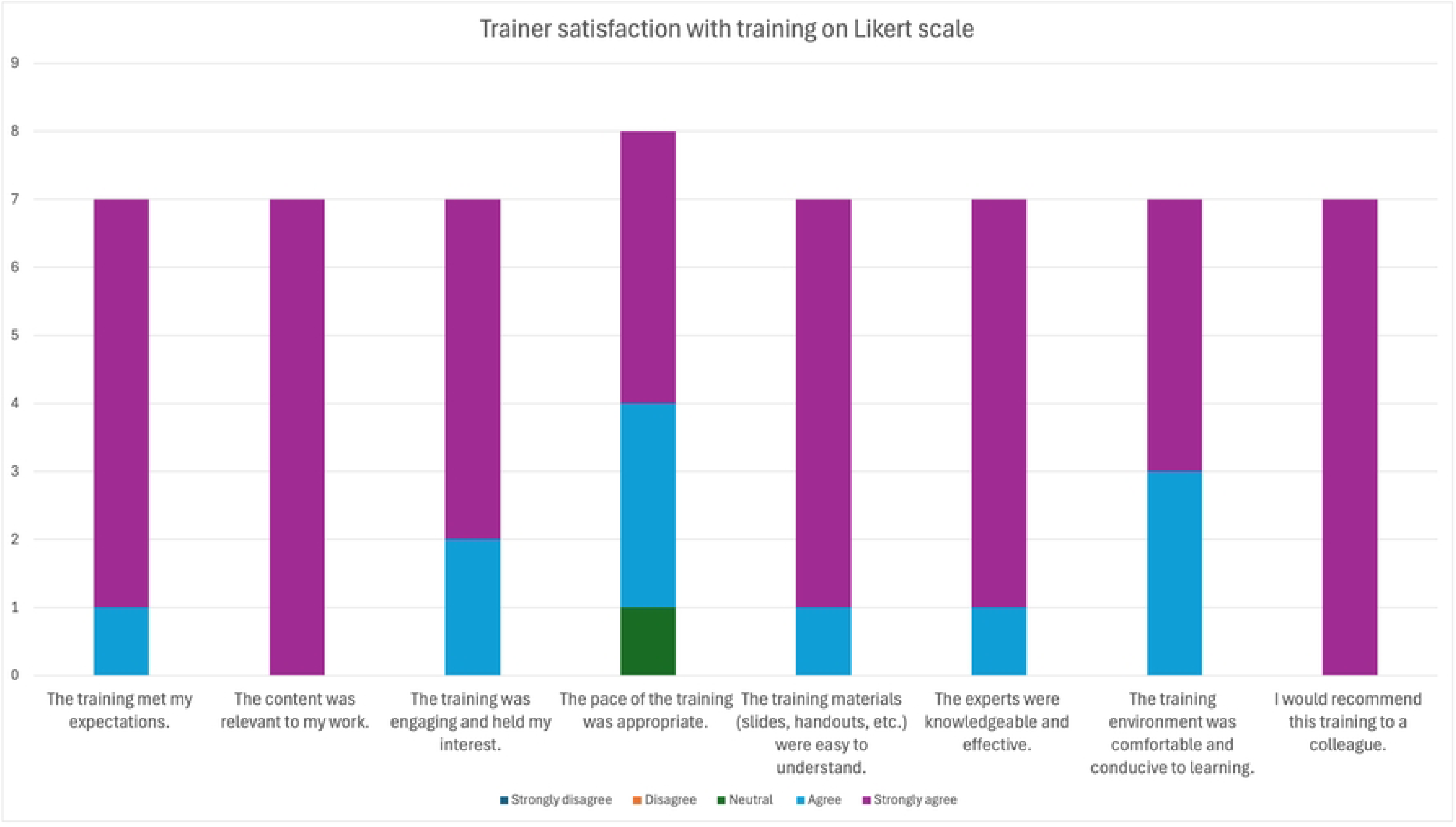
Trainer satisfaction with training on Likert scale

Master Trainers reported that the most valuable aspects of the echocardiography training were the opportunity for direct mentorship from expert cardiologists, learning structured pedagogical approaches, and gaining practical hands-on experience. Key challenges included limited time to cover the breadth of content, variation in baseline knowledge among trainees, and restricted opportunities for practice. Trainers described observing their trainees’ growth and increased competence as the most satisfying aspect of the program. To strengthen future iterations of the training, trainers recommended extending the duration of the program, improving pre-training preparation to standardize foundational knowledge, and providing ongoing mentorship and support after the training concluded. Recurring themes are detailed in Table 2.

**Table 2:** Themes from Level 1 evaluation [n=7] on trainer satisfaction. Only themes that arose three or more times are listed.

| Theme | Comments | Count |
| --- | --- | --- |
| Training Duration and content | • Training duration was too short<br>• Not enough hands-on time<br>• Trainers wanted additional training topics, material, and training | 9 |
| Satisfaction with training | • Appreciated presence of in-person experts<br>• Learning to teach a skill was a valuable aspect of training<br>• Learning practical skills was a valuable aspect of training<br>• Seeing trainees learn was satisfying | 6 |
| Challenges | • Trainers found it challenging to teach a skill without taking over from the trainee<br>• Challenging to teach trainees who had different levels of background knowledge | 5 |
| Suggested improvements to training | • Suggested evaluating trainee knowledge before and after training<br>• Challenging to teach trainees who had different levels of background knowledge<br>• Trainers suggested changes to the schedule for the week | 3 |

### Kirkpatrick Level 2 – expert evaluation of trainer competencies

All trainers scored Excellent or Very good in the ability to carry out cardiac POCUS and describe features of cardiac pathology to students. In one knowledge category of managing cases that were not straightforward, four trainers scored Very Good and three scored Good. In the teaching domain, most trainers scored Very Good and Good (n=6), with one scoring Excellent in their ability to give a didactic lecture and guide trainees to improve their POCUS image quality. Communication skills was the domain with the most variation in grading, though the majority of trainers (n=6) were graded Excellent and Very Good in the categories and one was graded as Good. Six trainers were scored as Excellent in the Participation and Attitude, domain with one trainer scoring good or fair in the six categories. This is depicted in Figure 7.

**Figure 7:**
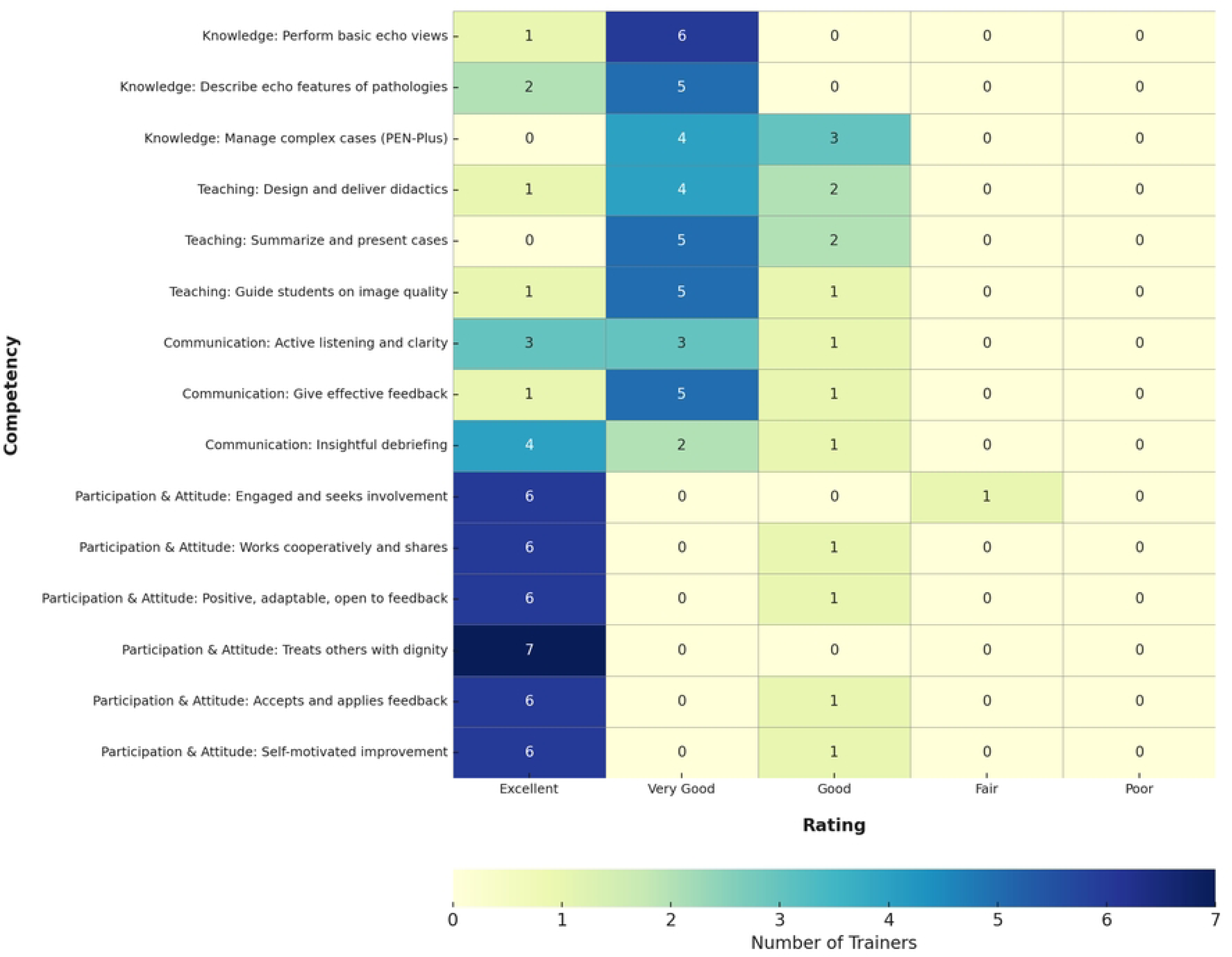
Expert evaluation of trainers by competency

### Level 2 – trainee evaluation of trainers

Most trainers scored highly when evaluated by their trainees with scores depicted in Figure 8.

**Figure 8:**
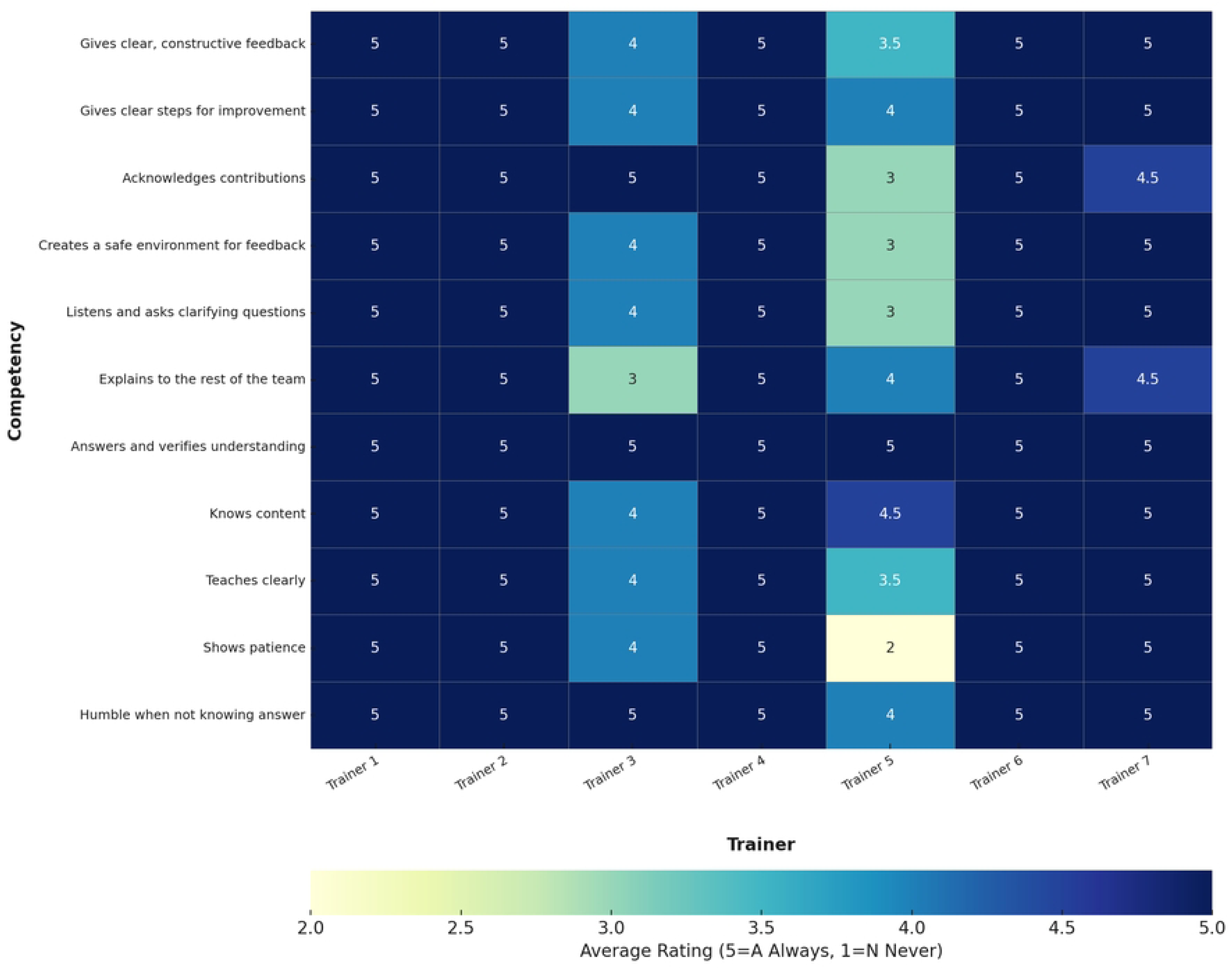
Trainer evaluations by their trainees

## Discussion

This paper presents a protocol for evaluation of a ToT program for cardiac POCUS among a cross-cultural group of trainers within the PEN-Plus strategy. Initial results from Level 1 and Level 2 of the Kirkpatrick model reveal high trainer satisfaction with the ToT program, good trainer performance when rated by experts, and good performance when rated by trainees with variation between trainers. Level 3 and Level 4 assessments will evaluate whether the ToT program changed trainer behavior and led to increased local cardiac POCUS training using the RE-AIM implementation framework as the evaluation tool. Correlations between immediate post-training evaluations (Kirkpatrick Level 1 and 2) and short-term implementation evaluations (Kirkpatrick Level 3 and 4) will be explored including whether expert and trainee assessments reflect short term implementation outcomes. Additional points of interest in the final analysis can explore whether the design of this ToT program is amenable to sustained results at 6 and 12 months.

Level 1 evaluations demonstrated that the ToT program as described in the methods was highly satisfactory: there was a structured curriculum, virtual training prepared trainers to perform supervised teaching during the in-person sessions, and in-person training practice supervised by experts was highly valued by the trainers. A “blended learning” approach incorporating both learning materials and interactive components, similar to our mixed virtual and in-person approach, has been found in other studies to be effective in adult learners including healthcare professionals, though the exact balance of each component is unclear (27). Level 1 evaluations in our study revealed that trainers wanted a longer duration, additional materials including well-defined objectives and competencies for the in-person week, and more hands-on time. Other studies have also underlined the importance of well-defined objectives and extending program time (27). Other elements in existing published data that have been found to predict successful ToT programs include support from Ministry of Health, context-appropriate training materials, and adequate resourcing and budgeting for training (14). We paid particular attention to these factors when planning our ToT program. One concern expressed in some studies is long-term sustainability given the high rate of staff turnover in many LMIC medical settings (27,28). Knowing this, we incorporated specific methodology for encouraging retention of the trainers in our ToT program, which was possible because of the overlap between the study team and the PEN-Plus programmatic team.

Level 2 evaluations of trainers by experts demonstrated that after training, there was still room for growth in knowledge and teaching in all trainers. The optimal duration and content of a cardiac POCUS ToT program may benefit from expansion by adding virtual sessions and an additional week of in-person training. This would also satisfy trainer feedback from Level 1.

Level 2 evaluations of trainers by trainees demonstrated a lower score for some trainers than others. While exact reasons for this were not captured in this study, cultural differences may have played a role. There is no existing literature to our knowledge in incorporating cross-cultural factors into ToT programs. While this study is not designed to determine whether trainee evaluations are associated with post-training implementation in trainers’ local clinics, exploring these associations during final data analysis may generate questions for further study.

There is published evidence that ToT models are effective in imparting skills and knowledge (14), significantly changing behavior (such as prescribing behavior) after training (27), and improving patient outcomes (27,29). A systematic review examining differing approaches to medical education in the field of stroke care found the ToT model in particular improved the quality of patient care in LMICs (29). A prospective ToT cohort study in antimicrobial resistance across 12 LMIC countries enabled trainees to take on future leadership and organizational roles [25,27,28]. PEN-Plus expansion relies on training of non-specialist providers in skills such as cardiac POCUS in order to make integrated care for severe chronic disease accessible to rural populations. PEN-Plus expansion is projected to occur exponentially as existing clinics open new clinics in additional regions in their countries, thus more trainers are needed. Decentralization of specific skills such as cardiac POCUS will likely rely on experienced non-specialist providers in the foreseeable future given the very low number of cardiologists in LMICs. Putting the RE-AIM evaluation in place gives trainers and clinics a well-defined and measurable way of tracking the effects of this and future ToT programs and emphasizes the importance of post-training follow-up. Training trainers who can also take on future leadership, administrative, and organizational roles within their PEN-Plus programs via ToT training would be a significant advantage of this approach.

Limitations of this study include the small sample size and single cohort of trainers. The response of trainers to the acceptability portion of the assessment (Kirkpatrick Level 1) may be biased by the pre-existing commitment of their sending organizations in using their skills for future training. The RE-AIM framework does not formally capture contextual barriers for training implementation, though the study team is aware of most barriers through programmatic work.

Taking into account existing studies, there is still very little that is known about variables that allow peer trainers in LMICs to carry out training effectively. Few ToT studies employ a standardized training evaluation tool such as the Kirkpatrick model or evaluate short- or long-term outcomes after training. This study adds to the body of literature on skills-based ToT programs in LMICs, and future studies incorporating implementation outcomes after ToT training would be valuable. Analysis of complete results from this multi-country ToT program will inform planning of future programs within the NCDI Poverty Network and will provide guidance for other training programs across a diverse range of LMICs.

## Author Contributions

All authors conceived the idea for the ToT program and participated in the methodology, design, and execution of the program. SLK drafted the original manuscript and remaining co-authors contributed to revisions to create the final version of the manuscript.

## Data Availability

All relevant data are within the manuscript and its Supporting Information files.

## Acknowledgements

The study team wish to thank Dr. Yvette Kisaka from the Ministry of Health in Kenya for supporting the in-person component of the ToT program as well as PEN-Plus implementation in Kenya alongside NCD Alliance Kenya. We thank Ramon Ruiz for assistance with formatting of the figures used in this paper.

---

Appendix 1: PEN-Plus adult heart failure management protocol.

Appendix 2: PEN-Plus pediatric heart failure management protocol

Appendix 3: Kirkpatrick Level 1, 2, and 3 Evaluations

